# How effective was Newfoundland & Labrador’s travel ban to prevent the spread of COVID-19? An agent-based analysis

**DOI:** 10.1101/2021.02.05.21251157

**Authors:** Dionne M. Aleman, Benjamin Z. Tham, Sean J. Wagner, Justin Semelhago, Asghar Mohammadi, Paul Price, Randy Giffen, Proton Rahman

## Abstract

**Background:** To prevent the spread of COVID-19 in Newfoundland & Labrador (NL), NL implemented a wide travel ban in May 2020. We estimate the effectiveness of this travel ban using a customized agent-based simulation (ABS).

**Methods:** We built an individual-level ABS to simulate the movements and behaviors of every member of the NL population, including arriving and departing travellers. The model considers individual properties (spatial location, age, comorbidities) and movements between environments, as well as age-based disease transmission with pre-symptomatic, symptomatic, and asymptomatic transmission rates. We examine low, medium, and high travel volume, traveller infection rates, and traveller quarantine compliance rates to determine the effect of travellers on COVID spread, and the ability of contact tracing to contain outbreaks.

**Results:** Infected travellers increased COVID cases by 2-52x (8-96x) times and peak hospitalizations by 2-49x (8-94x), with (without) contact tracing. Although contact tracing was highly effective at reducing spread, it was insufficient to stop outbreaks caused by travellers in even the best-case scenario, and the likelihood of exceeding contact tracing capacity was a concern in most scenarios. Quarantine compliance had only a small impact on COVID spread; travel volume and infection rate drove spread.

**Interpretation:** NL’s travel ban was likely a critically important intervention to prevent COVID spread. Even a small number of infected travellers can play a significant role in introducing new chains of transmission, resulting in exponential community spread and significant increases in hospitalizations, while outpacing contact tracing capabilities. With the presence of more transmissible variants, e.g., the UK variant, prevention of imported cases is even more critical.

## 1 Introduction

On March 11, 2020, WHO declared SARS-CoV-2 (COVID) a pandemic with 118,000 cases largely from four countries [1]; four months later, there were 28 million cases globally and 1.8 million new cases per week. [2]. Empirical evidence indicates that travel restrictions reduce the spread of diseases in general [3, 4], and successful current real-life strategies for COVID include travel restriction as a crucial component. In China, travel restrictions imposed on January 23, 2020 resulted in a decrease of 515 to 39 travel-related cases in Wuhan just one week later [5], and an estimated three-day delay of COVID arrival [6]. Continued travel restrictions in New Zealand, part of a broad intervention package, led to one of the lowest case numbers and mortalities worldwide [7] and effective community elimination of COVID by June 2020 [8]. Similarly, Australia utilized both international and internal travel restrictions, controlling both initial and subsequent waves [9].

Though seemingly effective at slowing COVID spread, the economic impact of travel restrictions is inconclusive, and mobility restrictions disproportionately impact poorer people [10] and workers in certain industries [11]. Further, travel restrictions may be legally controversial [12] and are only moderately effective unless implemented with other policies [13, 14]. Thus, it is important to understand the potential benefit of travel restrictions for individual regions.

For the province of Newfoundland & Labrador (NL) in Canada, strict restrictions on travel from out-of-province, implemented May 2020, have contributed to low case counts. NL has a large tourism economy, with annual non-resident travellers (≈533,000 [15]) exceeding the province’s population (≈520,000 [16]), as well as a large presence of rotational workers. Thus, there is considerable pressure to relax travel restrictions. While studies show that travel increases COVID spread, there is a reliance on models with “potentially inappropriate assumptions” [17]. We therefore use a granular agent-based simulation (ABS) of COVID spread in NL to estimate the effectiveness and appropriateness of NL’s travel ban, finding that a small number of infected travellers can introduce new chains of transmission even with contact tracing, similar to documented case studies [18, 19]. This finding is particularly important in the presence of new COVID variants of concern (VOCs)—particularly the UK variant, estimated to be 56% more transmissible than pre-existing variants [20]—as demonstrated by a recent outbreak at a long-term care home in Barrie, Ontario, where contact with an international traveller by a staff member quickly led to the infection of 127 residents and at least 32 deaths [21].

## 2 Methods

### 2.1 The agent-based simulation model

The ABS is adapted from morPOP (morLAB Pandemic Outbreak Planner), an ABS built to simulate the spread of pandemic influenza in the Greater Toronto Area, Ontario, Canada [22]. Specific adaptations for COVID include the delineation of pre-symptomatic (latent), symptomatic, and asymptomatic patient statuses. New features include travellers and contact tracing, while public transportation usage is removed as public transit is not heavily used in NL and little transit data was available. The population is constructed based on data from Statistics Canada’s 2016 census [16] and provincial departments, and households are located in census subdivisions (CSDs) according to the census; household compositions (single person, two adults, two adults with one child, etc.) are similarly determined by CSD-level census data.

morPOP models each individual in the population as a unique entity, allowing for individual behavioral, demographic, and health characteristics to be represented. Unique environments where people interact are represented: households, hospitals, schools, businesses, etc. Individuals have random community contacts in the CSDs where they live and work. Each individual has an infection status following the typical SIRD (susceptible, infectious, recovered, dead) model, with a latent asymptomatic infectious period after initial infection. During the simulation, individuals follow behavior patterns like being at home for a certain number of hours per day, going to work or school, observing physical distancing guidelines or not, and seeking medical care when infected. Visits to primary care physicians (assumed to be all in-person) and hospitals are captured, but use of advanced medical care (e.g., ventilators) is not.

At each location visited by an individual, there is interaction with other individuals at the same location (household members, fellow students, fellow employees, etc.), and an individual’s chance of becoming infected is determined by contact with infected individuals in each location and the nature of that contact. For example, contact in a household may be much more likely to result in infection than contact with a customer at a business. Individuals’ health statuses are updated each simulated day, and the probability that a susceptible person *j* becomes infected on day *n* is calculated by

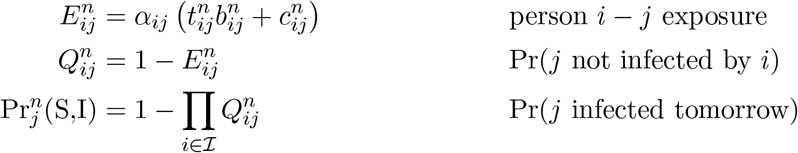

where *α*_*ij*_ is the Vancouver School of Economics (VSE) risk factor for the environment in which person *i* and person *j* have contact [23]; 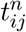 is the time (in minutes) of contact between *i* and *j* on day *n*; 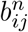 is the rate of disease transmission between *i* and *j* per minute of contact; 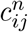 is indirect transmission between *i* and *j*; and *I* is the set of currently infected individuals.

Travellers arrive to CSDs either singly or in travel parties at a rate determined by monthly travel volumes, and each travel party (including a travel party of size one) forms its own household. The entire travel party is assigned to a workplace in the CSD if the visit purpose is business, or is “bubbled” with another household in the CSD if the visit purpose is to visit friends/family. For other visit reasons, the traveller (or travel party) only has random community contacts with residents. After a pre-determined length of stay, travellers leave the simulation. Infected travellers count towards the province’s infections while they are in the province, but not after they leave.

Specific limitations of the current implementation of morPOP for NL-COVID, and whether they are likely to result in an under- or overestimation of infections, are in Table 1.

**Table 1:** Limitations and potential bias of morPOP predictions.

| Limitation | Bias | Explanation |
| --- | --- | --- |
| Travellers for business assigned to random workplaces in their CSD | Under | Rotational workers are likely to work in the same workplaces (e.g., fish processing plant), significantly increasing the chance of infection for resident workers |
| Individual travellers only visit/work in one CSD | Under | Opportunity for disease transmission in multiple communities is not present |
| Transmission rates do not differentiate ages 5-18 | Over (?) | Although still uncertain, it is believed that younger children are less contagious than older children |
| Institutional facilities (long-term care homes, prisons) are not modelled | Under | Disease may spread rapidly in these environments |
| Transmission within hospitals not modelled | Under | Disease may spread in hospitals |
| Specific nature of workplace contact (indoor, outdoor, isolated offices/cubicles, etc.) not modelled | Over | All contacts and transmission within a workplace are assumed to happen like a face-to-face conversation, though VSE risk factors [23] are incorporated |

### 2.2 Model parameters

The main parameters for the ABS are shown in Table 2. Transmission rates are adapted from influenza per-minute age-based transmission rates [24]. Contacts in workplaces are adjusted by VSE Risk Factor [23, 24], and death rate is adjusted by age and comorbidity [25–27], where comorbidities are assigned to individuals by prevalence per CSD (publicly available Canadian Community Health Survey and Statistics Canada data). Hazard rates for identified comorbidities of interest (Table 3) are applied to both death and hospitalization probabilities. Demographic parameters for individuals and households are based on 2016 NL census data [16]. Workplace, home-work commutes, hospital, and Regional Health Authority parameters are from NL-specific data. As the purpose of this study is to examine the effectiveness of NL’s travel ban, transmission rates in non-household environments are scaled down by 80% to reflect protective behaviors (social distancing, masks) assumed to be commonplace during the first months of the pandemic.

**Table 2:** Model parameters.

| Parameter | Value |
| --- | --- |
| **Population** |  |
| Initial number of unknown infections in NL population | 5 |
| Duration of self-isolation | 14 days |
| Probability that a resident self-isolates upon symptom onset | 90% |
| Probability that an entire household self-isolates when one member becomes symptomatic | 90% |
| Number of days after symptom onset that a person chooses to alter behavior, with probabilities | 0 (80%), 1 (10%), 2 (10%) |
| Household bubble size | 2 |
| Daily age-based contacts outside home/work/school | S1 Dataset, Canada [28] |
| **Travellers** |  |
| Total annual non-resident travellers in a typical year (2018) | 533,507 [15] |
| Monthly non-resident travellers to each census subdivision (matched to economic zones) | Table 10 [29] |
| Travel party size, assumed to be of size 1-4 individuals and tuned to match known means | 1 (31%), 2 (63%), 3 (2%), 4 (4%) (Table 5 [29]) |
| Percent of travel parties who are assigned to a workplace (travel for business or conference) | 29% (Table 3 [29]) |
| Percent of travel parties who bubble with one household (travel for visiting relatives or other) | 36% (Table 3 [29]) |
| Length of stay (days) of non-resident parties, tuned so that the 32.1-day average stay for business travellers by auto is 4 standard deviations from the mean | Normal(10.8,5.3) (Table 6 [29]) |
| Number of days already been infected upon arrival for infected travellers (maximum possible is 5 days latent + 10 days symptomatic) | 0-15 days |
| **Contact tracing** |  |
| Daily contact tracing call capability | 1932 calls/day |
| Days after being infected that contact tracing begins (includes latent period, days with symptoms before getting tested, and turnaround time for test result) | 9 days |
| Number of days before symptom onset to contact trace | 2 days |
| Percent of non-medical-care seeking individuals that are contact-traced | 50% |
| Percent of known contacts that are successfully found and contacted | 95% |
| Percent of contacts outside home/work/school that an individual remembers | 90% |
| Number of employees for workplace to be small enough to be closed due to a single infection | 10 |
| **Disease** |  |
| Per-minute disease transmission rates between age groups | Table A1 [24] |
| Percent of asymptomatic cases | 42.5% [30] |
| Contagiousness of pre-symptomatic and asymptomatic cases compared to symptomatic | 25% [31] |
| Latent period | 5 days |
| Symptomatic period | 6-10 days |

**Table 3:** Hazard rates by comorbidity for death and hospitalization probabilities.

| Comorbidity | Hazard rate |
| --- | --- |
| Cancer | 2.05 |
| Chronic obstructive pulmonary disease (COPD) | 1.90 |
| Diabetes | 1.90 |
| High blood pressure | 1.58 |
| Ischemic heart disease (IHD) | 1.40 |
| Obesity | 1.40 |
| Asthma | 1.25 |

All traveller parameters (Figure 1) are determined from an exit survey conducted by the Government of Newfoundland and Labrador Department of Tourism, Culture, Industry and Innovation [29]. This exit survey indicates traveller destinations by economic zone, which were mapped to CSDs.

**Figure 1:**
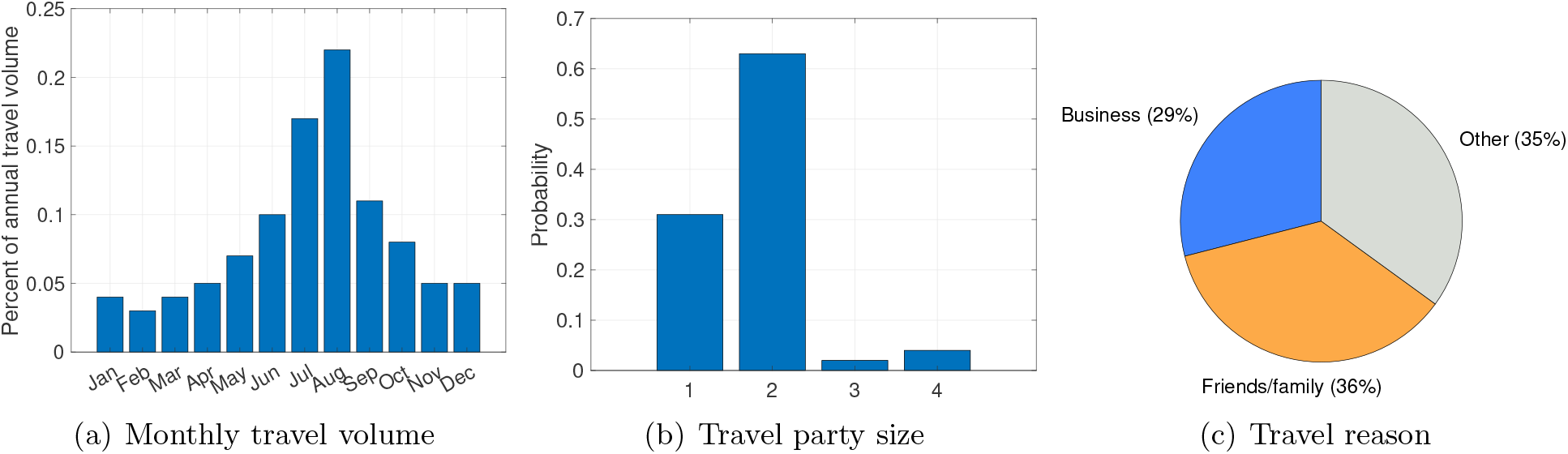
Traveller arrival details.

### 2.3 Travel scenarios

To assess the impact of the travel ban, we compare a baseline scenario of the travel ban in place with low, medium, and high values of travel volumes, traveller infection rates (*θ*), and traveller quarantine compliance rates (Table 4). The impact of contact tracing on each scenario is examined by running each scenario with and without contact tracing, for a total of 56 scenarios.

**Table 4:** Low, medium, and high values in traveller scenarios.

| | Travel volume* | Traveller infection rate ($\theta$) | Traveller quarantine compliance rates (immediately / after symptom onset) |
| --- | --- | --- | --- |
| Baseline | 0% (travel ban) | – | – |
| Low | 24% | 0.03% | 50% / 25% |
| Medium | 50% | 0.1% | 75% / 50% |
| High | 100% | 1% | 80% / 80% |

\* Travel volume is in terms of non-resident travel volume; 24% non-resident travel volume is approximately 10% total travel volume (non-resident + resident).

A percentage of travellers (50%, 75%, 80%) immediately quarantine upon arrival, meaning that they only have contacts within their household (travel party) for the duration of the quarantine. Of the travellers who did not quarantine, infected individuals will choose to begin quarantine upon symptom onset with a certain probability (25%, 50%, 80%). These lower probabilities reflect the intuition that travellers who have already chosen to not follow quarantine rules are likely incentivized to continue not following those rules when symptoms appear due to the nature of their travel (strict timelines, work/family pressures) and demonstrated lack of adherence to quarantine rules.

Reasonable low (0.03%), medium (0.1%), and high (1%) values for *θ* were chosen in consultation with NL Public Health, noting that a 1% infection rate was calculated at Toronto Pearson International Airport from September to November 2020 [32]. The medium (50%) and high (100%) travel volumes were similarly estimated at the time the travel ban was implemented. After the summer, St. John’s International Airport in NL reported that travel volumes were in fact 10% of typical volumes. As the available travel volume data [29] is for non-resident travel only, 10% of total travel volumes is approximately 24% non-resident travel volumes, and thus, 24% travel volume is examined as the low travel volume scenario.

The scenarios start on May 1 and simulate 100 days. An initial undetected five cases are present at the start of the simulation. Although NL had no detected cases on May 1, without full-scale population testing, it is reasonable to assume that there are a small number of existing infections that are asymptomatic or have sufficiently mild symptoms so as not to require hospitalization. Households are engaged a “double bubble”, meaning that households have close contact with one other household, in accordance with NL’s de-escalation protocol during the time period.

### 2.4 Model implementation

Unlike most ABSs that struggle to capture large populations or that require lengthy computation times and memory, morPOP is written in C++ for computational speed and parallelization. Using a number of computational techniques specifically designed to speed up run times, morPOP simulated a single 100-day outbreak on the NL population of ≈ 520, 000 agents in on average 1.8min without travellers or contact tracing; 5min with travellers; and 8.3min with travellers and contact tracing. The difference in time is primarily due to dynamic memory allocation required by travellers and contact tracing. The model was parallelized with one simulation per processor and implemented on high-performance computing infrastructure provided by the Center for Health Informatics and Analytics (CHIA) at Memorial University. The specific infrastructure used was three Linux nodes, each with 32 cores (64 threads) and 256 GB RAM, allowing for 192 simulations to be run simultaneously. Thus, a run of 500 simulations requires ≈5-25 min, depending on the presence of travellers and contact tracing. are applied to both death and hospitalization probabilities. Demographic parameters for individuals and households are based on 2016 NL census data [16]. Workplace, home-work commutes, hospital, and Regional Health Authority parameters are from NL-specific data. As the purpose of this study is to examine the effectiveness of NL’s travel ban, transmission rates in non-household environments are scaled down by 80% to reflect protective behaviors (social distancing, masks) assumed to be commonplace during the first months of the pandemic.

## 3 Results

Five hundred simulations were performed for each scenario. To assess the impact of contact tracing, we first analyze scenarios without contact tracing, followed by the same scenarios with contact tracing.

### 3.2 Without contact tracing

In the baseline (travel ban) scenario, the number of infections reaches 0 in late June, while all other scenarios show exponential increase (Figure 2, top). Travellers’ quarantine compliance does not have a large impact on community disease spread; once infected travellers initiate community disease spread, exponential growth of infections begins. Travel volumes, however, significantly impact disease spread, with 1% infected travellers yielding such high numbers of cases that the figure had to be plotted on a different scale. Table 5 indicates the magnitude increase in mean total cases and hospitalizations over baseline for each scenario, again illustrating that quarantine compliance impacts total cases far less than travel volume and infection rate.

**Table 5:** Magnitude increase over baseline (travel ban) scenario, without contact tracing. Ranges reflect differences due to traveller quarantine compliance.

| Travel volume | Mean total cases |  |  | Mean peak hospitalizations |  |  |
| --- | --- | --- | --- | --- | --- | --- |
| | $\theta = 0.03\%$ | $\theta = 0.1\%$ | $\theta = 1\%$ | $\theta = 0.03\%$ | $\theta = 0.1\%$ | $\theta = 1\%$ |
| 100% | 14-18x | 20-24x | 80-96x | 14-18x | 20-25x | 77-94x |
| 50% | 10-12x | 18-19x | 52-60x | 10-12x | 17-20x | 53-64x |
| 24 % | 8-11x | 12-14x | 28-33x | 8-10x | 12-14x | 29-35x |

**Figure 2:**
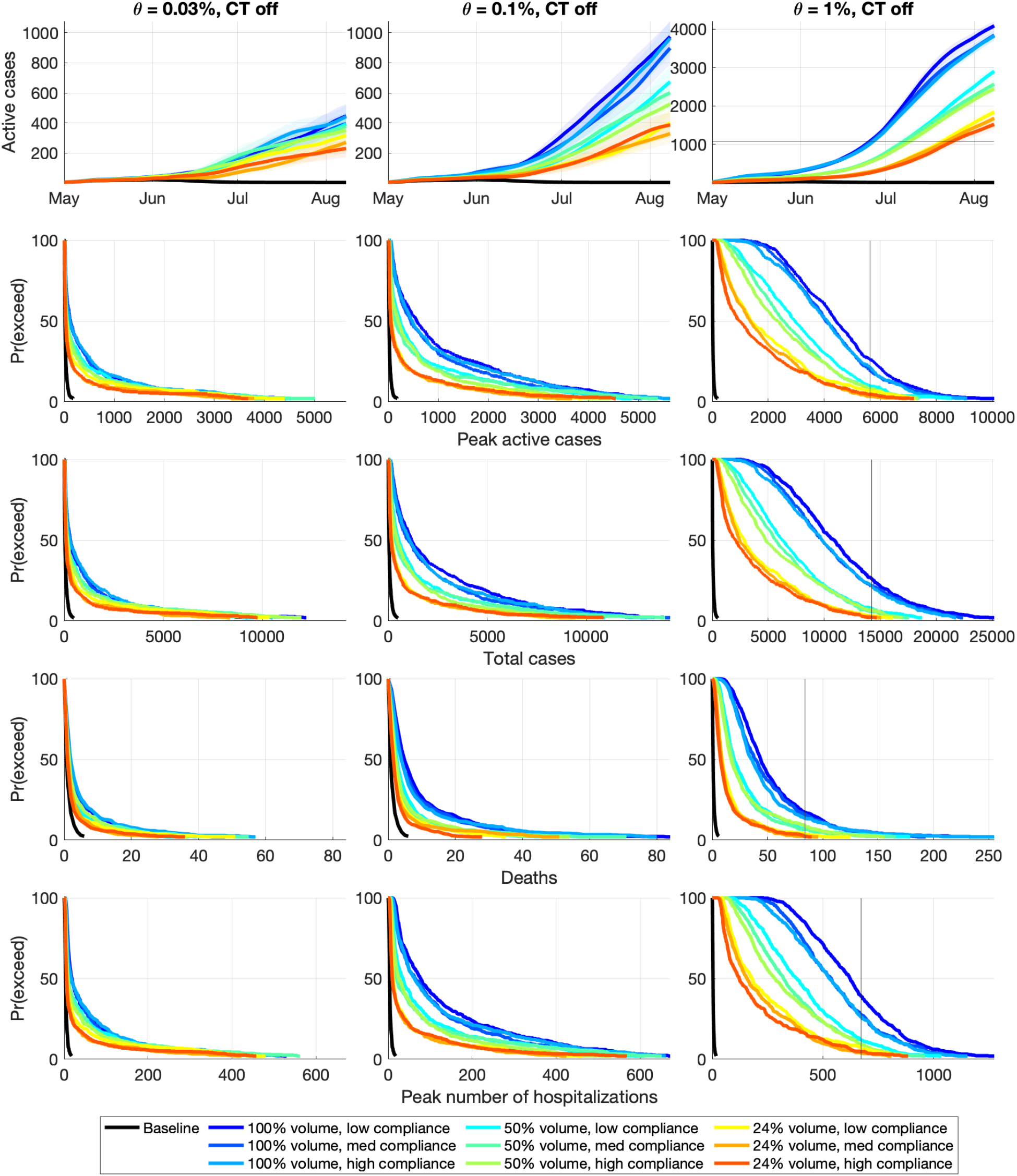
Simulation outcomes without contract tracing (CT). 95% confidence intervals are shown, and the mean is bolded. *θ* is the traveller infection rate. Note that *θ* = 1% is on a different scale; the horizontal line at ≈800 cases in *θ* = 1% active cases (top) indicates the upper limit of the figures for *θ* = 0.03% and 0.1%, while the vertical lines in other rows indicate the maximum *x*-axis value for *θ* = 0.03% and 0.1%.

Because mean and confidence interval plots like those in active case plots of Figure 2 (top) obscure simulation outcomes that are much larger than the mean (e.g., super-spreader events), simulation outcomes are better illustrated by examining the probability that metrics exceed thresholds. The other rows in Figure 2 illustrate the probability that the peak number of cases, total cases, deaths, and hospitalizations exceeds the threshold in the *x*-axis. In all metrics, for a fixed rate of infection, the probability of increasingly worse outcomes increases as the travel volume increases and rate of compliance declines. As the rate of infection increases, the outcomes all worsen dramatically, particularly as the rate of infection grows to 1%.

### 3.2 With contact tracing

Contact tracing is highly effective at reducing disease spread (Figure 3), but the predictions of daily active COVID cases still exhibit exponential growth even in the best-case scenario of 24% travel volume, *θ* = 0.03%, and high quarantine compliance. The number of cases is still determined much more by travel volume and traveller infection rate rather than compliance, and contact tracing is insufficient to stop community spread from travellers (Table 6), which also results in significantly more hospitalizations than with the travel ban in place.

**Table 6:** Magnitude increase over baseline (travel ban) scenario, with contact tracing. Ranges reflect differences due to traveller quarantine compliance.

| Travel volume | Mean total cases |  |  | Mean peak hospitalizations |  |  |
| --- | --- | --- | --- | --- | --- | --- |
| | $\theta = 0.03\%$ | $\theta = 0.1\%$ | $\theta = 1\%$ | $\theta = 0.03\%$ | $\theta = 0.1\%$ | $\theta = 1\%$ |
| 100% | 3-4x | 6-7x | 46-52x | 3-4x | 6-7x | 43-49x |
| 50% | 3-4x | 4x | 23-26x | 3x | 4x | 22-25x |
| 24% | 2-3x | 3-4x | 12-13x | 2x | 3x | 12-13x |

**Figure 3:**
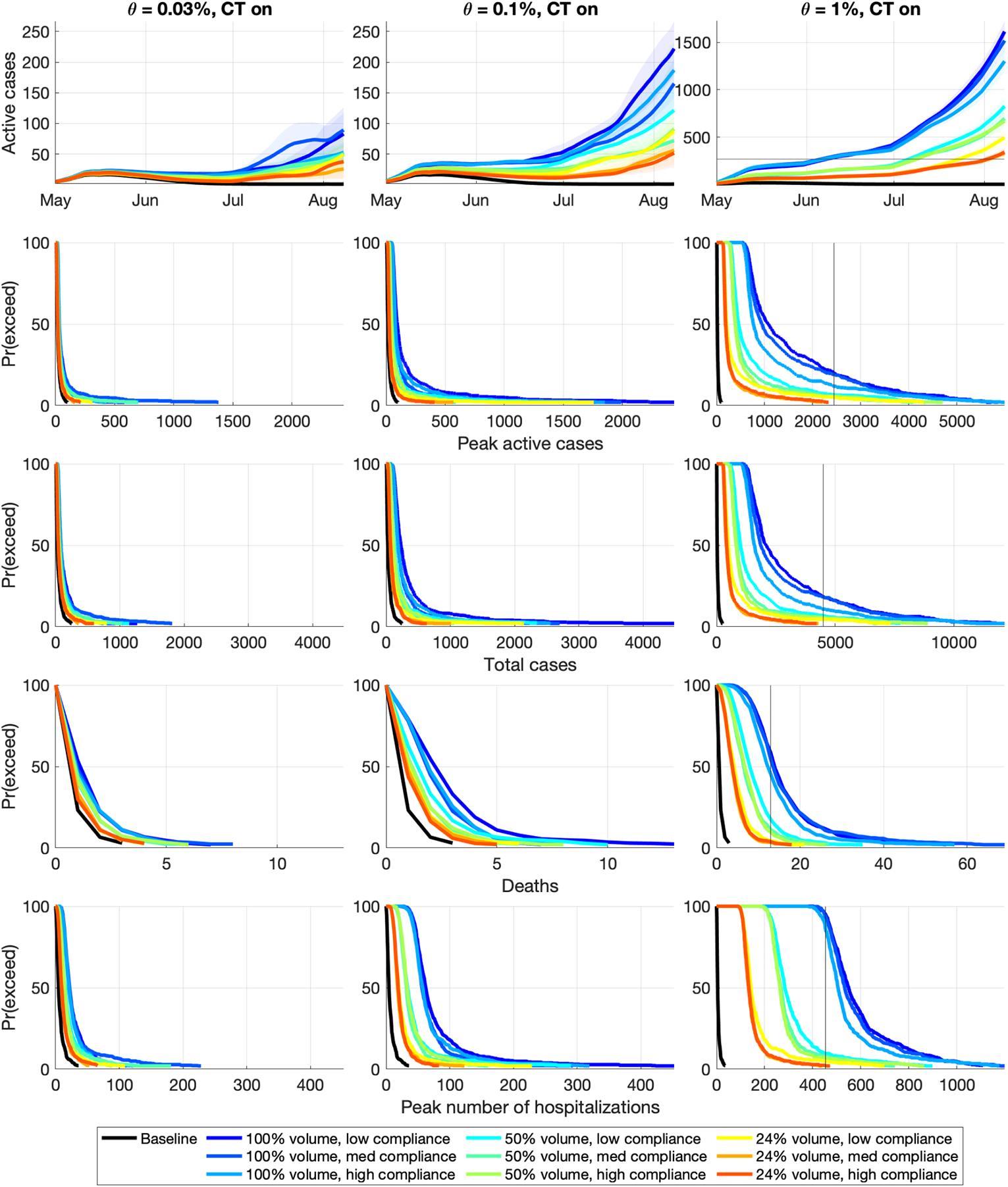
Simulation outcomes with contract tracing (CT). 95% confidence intervals are shown, and the mean is bolded. *θ* is the traveller infection rate. Note that *θ* = 1% is on a different scale; the horizontal line at ≈140 cases in *θ* = 1% active cases (top) indicates the upper limit of the figures for *θ* = 0.03% and 0.1%, while the vertical lines in other rows indicate the maximum *x*-axis value for *θ* = 0.03% and 0.1%.

Contact tracing reduces the mean total number of cases by 45.5% in the baseline scenario, and is effective in the presence of travellers (though still unable to stop exponential growth), but effectiveness decreases as the rate of infected travellers increases. At *θ* = 0.03% and *θ* = 0.1%, contact tracing reduces the mean total number of cases by 81.4-89.6%, and by 67.6-79.3% for *θ* = 1%.

The daily call capacity for NL is 1932 calls, which has about a 10% and 16% chance of being exceeded in the worst-case travel volume and compliance scenarios for *θ* = 0.03% and *θ* = 0.1%, respectively (Figure 4). For *θ* = 1%, the best-case travel volume scenario has call overload around 15-23% likelihood; the other scenarios have a 37-72% chance of call overload, and a 21-45% chance of even exceeding 5000 calls per day.

**Figure 4:**
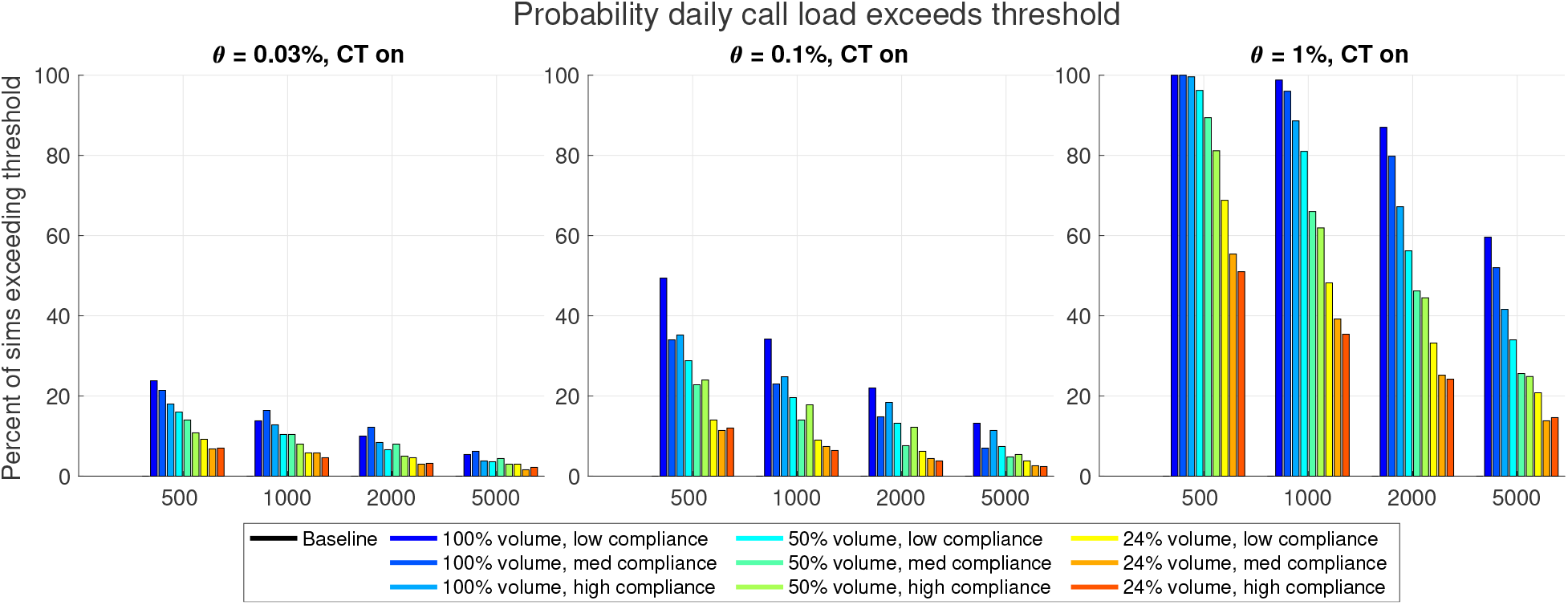
Contact tracing daily call load

## 4 Interpretations

If the existing number of cases in NL is very small as was believed at the time of the travel ban, even a very small number of infected travellers had the potential to dramatically worsen the pandemic even with high levels of traveller quarantine compliance. However, the mere act of travel, especially by air (which comprises 84% of NL’s non-resident travel [29]), increases exposure to COVID, and travellers may therefore have a higher risk of infection than the general population of their province or country of origin. Contact tracing has a tremendous ability to reduce the number of cases in the community, but even the best-case scenario of travellers begins exponential growth before the end of the 100-day simulation period. In all scenarios with travellers, both the mean total cases and peak number of hospitalizations were orders of magnitude larger than with the travel ban in place. The likelihood of exceeding contact tracing capacity is a concern even for lower rates of traveller infection at full travel volume, and capacity is very likely to be exceeded at a 1% rate of infected travellers.

Travel restrictions may be lessened with screening, however, allowing more travel means an increase in imported cases given the potential for false negatives [33–35], especially for asymptomatic and pre-symptomatic cases that may comprise close to half of infected travellers [36]. As demonstrated, the potential for a failure to contain COVID is high even with few infected travellers in the community, so any policy to reduce quarantine requirements must be carefully investigated with detailed knowledge of travellers’ points of origin and likely infection rates; however, such information is difficult to ascertain given potentially convoluted travel paths.

Thus, the morPOP simulation model indicates that the travel ban provided significant protection to the NL population. With more transmissible VOCs recently discovered, the importance of preventing imported cases from travellers is even more critical than the model results suggest.

## Data Availability

Some publicly available population data

